# Heavy Metals Induced Health Risk Assessment Through Consumption of Selected Commercially Available Spices in Noakhali District of Bangladesh

**DOI:** 10.1101/2023.02.06.23285555

**Authors:** Md Shahedul Islam, Akibul Islam Chowdhury, Lincon Chandra Shill, Sompa Reza, Mohammad Rahanur Alam

**Affiliations:** Department of Food Technology and Nutrition Science, Noakhali Science and Technology University, Noakhali, Bangladesh; Institute of Nutrition and Food Science, University of Dhaka, Dhaka, Bangladesh

**Keywords:** Food safety, Heavy metals, Spices, Atomic Absorption Spectrophotometry, Health Risk Assessment

## Abstract

There are growing concerns for food safety due to the risks associated with heavy metal contamination of culinary herbs and spices in developing countries like Bangladesh. The objective of the present cross-sectional study is to determine the concentrations of the heavy metals Lead (Pb), Cadmium (Cd), Chromium (Cr), Copper (Cu), and Iron (Fe) in the branded and non-branded spices collected from the Noakhali district by Atomic absorption spectrophotometry method, as well as to assess the health hazard risk associated with heavy metal intake via consumption of spices. The findings revealed that the greatest concentrations of Pb (15.47 ± 1.93), Cd (1.65 ± 0.011), Cr (31.99 ± 3.97), Cu (18.84 ± 1.97), and Fe (9.29 ± 1.71) were found in Cardamom, Coriander leaf, Bay leaf, Dried chili, and Black pepper respectively. Around 37% of Cr and 5% of Fe Estimated Daily Values (EDI) were greater than reference doses (RfD). All spices had Total Hazard Quotient (THQ) values for Pb, Cd, Cu, and Fe that were below acceptable, and 37% of all spices had TTHQ values for Pb, Cd, Cu, and Fe that were over the standard range, suggesting adverse health impacts for consumers. Green chili, ginger, coriander leaf, and all kinds of chili powder and turmeric powder have been reported to have exceptionally high TTHQ levels of Cr. The estimated carcinogenic risk for unbranded chili, green chili, and coriander leaf was found to be higher than safe levels.

## Introduction

Globally, food safety is a top priority for the public. Researchers have focused on the hazards associated with consuming contaminated foodstuffs, such as pesticides, heavy metals, or toxins in vegetables and spices, in response to the rising demand for food and food safety (1). Due to their abilities as a taste enhancer, color imparter, texture developer/improver, palatability, and medicinal effects, spices are natural ingredients in nearly all traditional cuisines around the world (2-4). More than a hundred distinct spices are grown across the world, with Asia being the primary producer (5). The global trade of spices is around USD 3 billion, while the reported per capita daily intake of spices is around 5 g (6).

Heavy metals are one of the severe pollutants in the environment due to their toxicity, persistence, and bioaccumulation problem. Heavy metal refers to heavy metallic chemical elements that have relatively high density and are toxic or poisonous at low concentrations (7). They include essential and non-essential elements such as Mercury, Cadmium, Chromium, Copper, Zinc, Manganese, Lead, etc. These heavy metals come in contact with over bodies via food, drinking water, and air. They are unsafe as they tend to bioaccumulate (increase in concentration in biological cells over time) (8). The toxicity can cause a variety of health problems, including kidney diseases, neurobehavioral and developmental abnormalities, high blood pressure, and cancer (9, 10).

Human exposure to harmful heavy metals has grown as a result of rising populations, which places greater strains on infrastructure and buildings. Soils that may be utilized to raise crops are contaminated by industrial spills and inadequate handling of chemical waste (11). Human activities such as mining, fertilizer-based agriculture (including aquaculture), and chemical fishing also lead to the transport and deposition of heavy metals in soils and water (12).

Global concern is growing regarding the widespread contamination of culinary herbs and spices such as curry powder, oregano, black pepper, and turmeric with heavy metals such as lead.(13). As a matter of fact, spices are one of the top five food items that are most frequently contaminated (14, 15). Markets in the United States, Europe, and other regions with well-resourced regulatory authorities nevertheless report cases of contamination of culinary herbs and spices, despite strict regulation.. Recently, excessive amounts of lead were found in several brands of turmeric sold in the United States, drawing attention to the potential health risks posed by consuming certain culinary herbs and spices. (16). The consequences of investigations by the FDA caused these turmeric brands to be removed from the shelves (13). Regulatory organizations in developing countries such as Bangladesh do not sample spices on the market on a regular basis to assess their safety, maybe because these heavy metals are not deemed a hazard. Very little research has been directed to evaluate heavy metal-related risk assessment in spices in Bangladesh. Rahman, M.A. et al. found twice the amount above the acceptable value of lead in unbranded turmeric samples collected from Dhaka city using atomic absorption spectroscopy (17). Akter, S., et al found similar result using the Ion Beam Analysis Technique (18). There is no other study has been conducted to determine the heavy metal content and associated health risk through the consumption of spices in the coastal Noakhali region of Bangladesh. Being a coastal area, Noakhali is more prone to heavy metal accumulation and subsequently to human health risks (19).

Our present study focuses on determining the concentrations of the heavy metals Lead, Cadmium, Chromium, Copper, and Iron in the branded and non-branded spices collected from the Noakhali district. as well as assessing health hazard risk associated with heavy metal intake through spices consumption.

## Methods and Materials

### Study design and Sampling

Our current study is a cross-sectional study. A multi-stage probability sampling method was used to collect data on spice consumption frequency from 180 households. A household survey was conducted in six different clusters from three stratified locations. There were two small towns, two paurashavas, and two unions among these six. The six clusters are marked in **Figure 1**, and these are identified as Chowmuhani, Maijdee, Bashurhat, Kabirhat, Noakhali Union, and Underchar Union.

**Figure 1.**
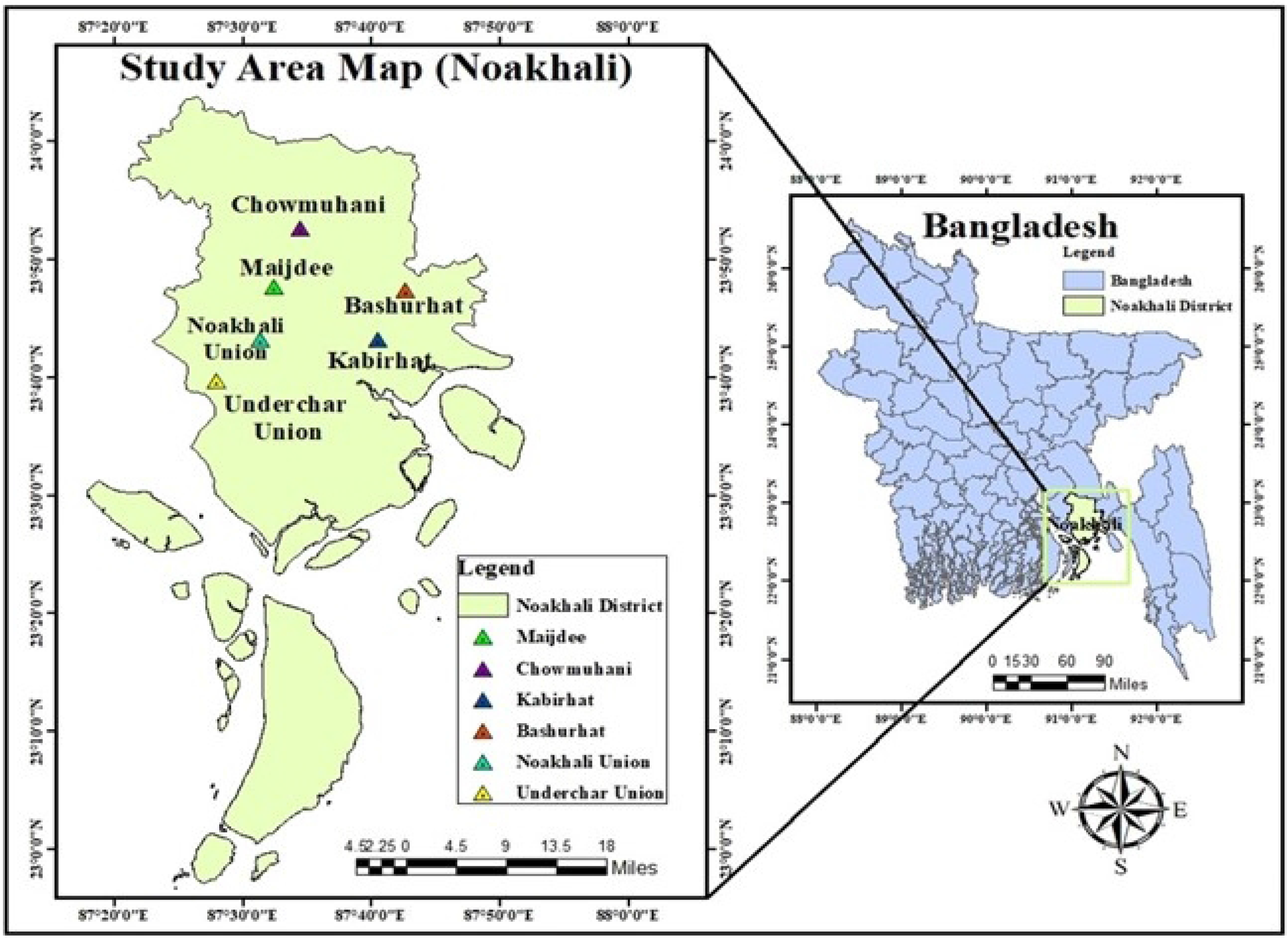
The map shows selected clusters in the Noakhali District of Bangladesh

### Data collection

Respondents were asked a set of relevant questions using a structured questionnaire with the remaining three sections, which included household information, demographic & anthropometric information, and consumption frequency of spices.

### Exclusion or inclusion criteria

Only those families whose heads gave their consent to participate in the study had their data collected. Anthropometric data were also omitted for members of the family who did not ingest spice-containing foods.

### Sample Collection and Preparation Sample collection

Nineteen samples of spices in commercially available sizes (50–125g) were taken from Noakhali’s Sonapur and Maijdee marketplaces. Non-branded spice samples (n = 15) and branded spice samples (n = 4) were acquired at random from open markets (Table 1).

**Table 1.** Classification of spice samples under research.

| **Sample ID** | **Local Name** | **English Name** | **Nature** | **Scientific Name** |
| --- | --- | --- | --- | --- |
| S-1 | Morichher<br>Gura | Chili Powder | Non-Branded | *Capsicum annuum L* |
| S-2 | Holuder Gura | Turmeric Powder | Non-Branded | *Curcuma longa* |
| S-3 | Morichher<br>Gura | Chili Powder | Branded | *Capsicum annuum L* |
| S-4 | Holuder Gura | Turmeric Powder | Branded | *Curcuma longa* |
| S-5 | Kancha<br>Morich | Green Chili | Non-Branded | *Capsicum annuum* |
| S-6 | Shukna<br>Morich | Dried Chili | Non-Branded | *Capsicum annuum* |
| S-7 | Elachi | Cardamom | Non-Branded | *Elettaria cardamomum* |
| S-8 | Garam Masala | Masala Mix | Branded | - |
| S-9 | Lobongho | Cloves | Non-Branded | *Syzygium aromaticum* |
| S-10 | Dhoniya Gura | Coriander Powder | Non-Branded | *Coriandrum sativum. L.* |
| S-11 | Aadha | Ginger | Non-Branded | *Zingiber officinale* |
| S-12 | Darucini | Cinnamon | Non-Branded | *Cinnamomum verum* |
| S-13 | Methi Pata | Fenugreek | Non-Branded | *Trigonella foenum-graecum* |
| S-14 | Jirar Gura | Cumin | Non-Branded | *Cuminum cyminum* |
| S-15 | Jira o Dhoniya<br>Mix | Cumin & Coriander<br>Mix | Non-Branded | - |
| S-16 | Jirar Gura | Cumin | Branded | *Cuminum cyminum* |
| S-17 | Dhoniya Pata | Coriander Leaf | Non-Branded | *Coriandrum sativum* |
| S-18 | Tej Pata | Bay Leaf | Non-Branded | *Laurus nobilis* |
| S-19 | Gool Morich | Black Pepper | Non-Branded | *Piper nigrum* |

### Wet Digestion of Samples

Wet digestion was carried out using a 1:4 concentrated mixture of HNO_3_ and H_2_S0_4_. The mixture was heated to 130°C using a thermostat-controlled heating block for one hour and was allowed to cool gradually. Each digestion mixture received 2 ml of H_2_O_2_, and the contents were returned to the oven for further heating. This step was replicated unless a colorless solution was generated. Following filtration, the clear solution was transferred to a 50 ml volumetric flask tube and topped up to the maximum with deionized water.

### Heavy Metal Determination with Atomic Absorption Spectroscopy

The heavy metal determination was performed with a PerkinElmer Inc. PinAAcleTM 900H Atomic Absorption Spectrometer (AAS) with dual beam optics continuum source double-beam background correction using a high-intensity deuterium arc lamp. The equipment was calibrated with a linear calibration through zero using a standard solution and a blank solution containing known concentrations of the heavy metals under investigation. The AAS was performed in triplicate on digested clear solutions for each sampled spice. The linearity (R2) ranges from 0.9897 to 0.9983 for the standards. The limit of detection (LOD) was set at 0.01 ppm/mL, and the standard measuring time for accurate determination was 3 seconds (20).

### Human Health Risk Assessment

An Estimated Daily Intake (EDI) is commonly used to determine the estimated health risk associated with consuming heavy metal contaminant (21).

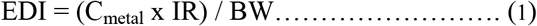

Where, C_metal_ implies the concentration of the metals in mg/kg, IR or ingestion rate expressed in gram per day per person, and BW is average body weight of the household members in kg.

Total Hazard Quotient (THQ) was used to assess the non-carcinogenic risks associated with long-term heavy metal exposure. Total Hazard Quotient (THQ) is the ratio of a substance’s levels of exposure over such a specific duration to a reference dose (RfD). Exposure levels below a certain level (<1) indicate a lower risk of adverse health effects. Consumption of tainted spices, on the other hand, may cause health hazards if the calculated value of THQ is equal to or greater than 1 (≥ 1) (22).

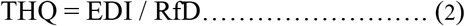

The RfD values for Pb, Cd, Cr, Cu, and Fe are 0.0035, 0.0005, 0.0003, 0.040, and 0.007 mg per kg per day, respectively (23-25).

The Total Target Hazard Quotient (TTHQ) was used to estimate the overall non-carcinogenic risk to human health from exposure to various heavy metals. THQ is the ratio of a substance’s levels of exposure over such a specific duration to a reference dose (RfD). Exposure below a certain level (<1) indicates a lower risk of adverse health effects. Consumption of tainted spices, on the other hand, may cause health hazards if the calculated value of THQ is equal to or greater than 1 (≥ 1) (22). The population may experience adverse health effects if the TTHQ value exceeds 1 (26-28).

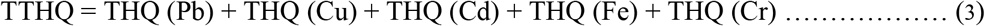

Carcinogenesis risk refers to an individual’s increased likelihood of developing cancer due to contact with hazardous carcinogenic compounds such as Pb, Cd, and Cr (29).

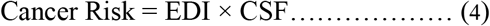

Where CSF (Cancer Slope Factor) is presented in mg/kg/day

The US Environmental Protection Agency recognizes a cancer risk in the range of 1 × 10^−6^ to 1 × 10^−4^ as appropriate for risk management purposes (30).

## Result and Discussion

### Concentrations of Selected Heavy Metals in Sampled Spices

**Figure 2(a-e)** summarizes the concentrations of selected heavy metals, *i*.*e*., Pb, Cd, Cr, Cu, and Fe, in the samples selected for this study. It was discovered that the levels of heavy metals in various spices varied substantially. The findings of this investigation are compared to international regulatory requirements and published literature.

**Figure 2.**
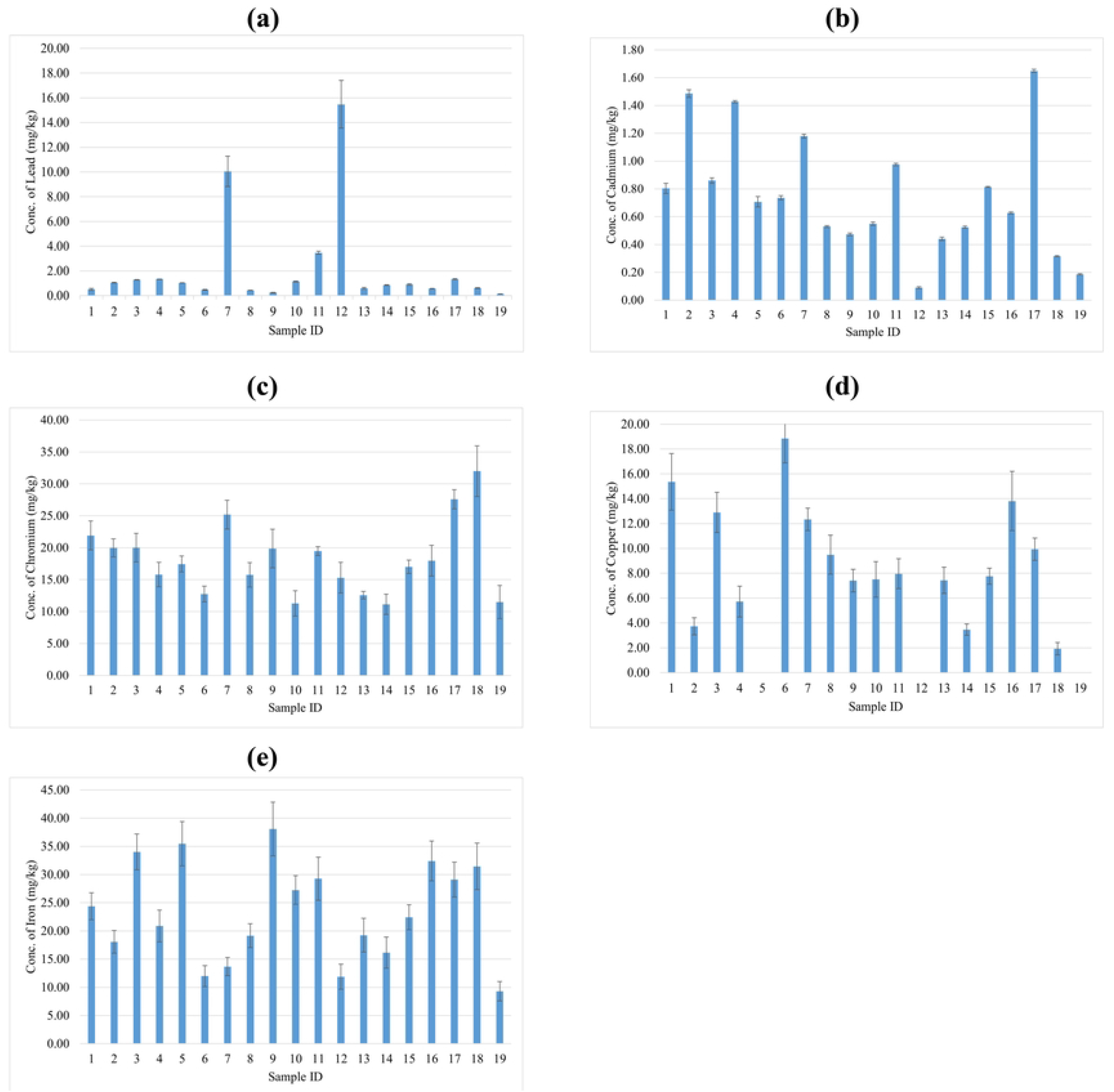
Mean concentration of heavy metals: (a) Lead (Pb), (b) Cadmium (Cd), (c) Chromium (Cr), (d) Copper (Cu), and (e) Iron (Fe) in sampled spices (mg/kg)

Lead (Pb) is considered as a non-essential level, and it mainly damages the kidneys and liver of the body (31). Cardamom had the highest Pb content (15.47 ± 1.93) mg/kg, followed by cinnamon (10.05 ± 1.22) mg/kg (Figure 2a). When compared to studies done in other countries, we find that concentrations in Pakistan, Romania, and Iraq, respectively, ranged from 4.44–15.88 mg/kg (22), 0.04–1.28 mg/kg (32), and 3.21–6.98 mg/kg (33). Chili powder has been shown to contain the highest levels of lead in Pakistan and Iraq (22, 33).

Consumption of cadmium can cause severe diseases even at low concentrations by accumulating in the kidneys and liver (34, 35). The highest concentration of Cd was found in coriander leaf (1.65 ± 0.01) mg/kg, non-branded turmeric powder (1.49 ± 0.02) mg/kg, and branded turmeric powder (1.43 ± 0.01) mg/kg (Figure 2b). Most Cd was found in Garam Masala, coriander, and curry powder in Pakistan, South Korea, and Iraq, respectively (22, 33, 36).

A high concentration of Cr can cause damage to the kidney, liver, and blood cells, although its deficiency causes hyperglycemia and elevated fat levels (35). Among the studied spices, we found that the greatest Cr levels were found in bay leaf (31.99 ± 3.97) mg/kg, coriander leaf (27.58 ± 1.50) mg/kg, and cardamom (25.19 ± 2.26) mg/kg (Figure 2c). In Ethiopia (37) and Bangladesh (38), the uppermost Cr concentration was discovered in coriander. In Saudi Arabia (39) and Iraq (33), it was found in cinnamon and curry powder, respectively. Copper (Cu) is an important trace element that plays a vital role in some biological processes of the human body; however, at high concentrations, it can damage the kidneys and blood cells (40).

The most significant Cu concentrations were found in dried chili (18.84 ± 1.97) mg/kg, non-branded chili powder (15.36 ± 2.28) mg/kg, and branded cumin (13.81 ± 2.38) mg/kg, respectively (Figure 2d). The highest copper level was found in black pepper in Saudi Arabia(39), Turkey (41), and Iraq (33), as well as in turmeric powder in Bangladesh (38).

Black pepper had the greatest iron content, at 9.29 ± 1.72 mg/kg, followed by green chili and branded chili powder, with concentrations of 35.46 ± 3.94 mg/kg and 34.02 ± 3.18 mg/kg, respectively (Figure 2e). Compared to comparable research conducted in Iraq, it was shown that the ranges of Red pepper (791.66 ± 6.57) mg/kg and curry powder (830.091 ± 11.54) mg/kg had the greatest quantities of elemental Iron (41).

### Estimated Daily Intake (EDI) of Selected Heavy Metals

The estimated daily intake of heavy metals from spices is presented in Table 2. EDI of Cr in some spices such as chili Powder (NB), Turmeric Powder (NB), Chili Powder (B), Turmeric Powder (B), Green Chili, ginger, and coriander leaf exceeded the reference dose (RfD) levels; however,,, none of the heavy metals exceeded the maximum permissible limits (MPL). The EDI of heavy metals through consumption of spices were in order of Fe>Cr>Cu>Pb>Cd. A study discovered an elevated EDI of Cr (0.070) for turmeric (42). Due to the absence of permissible tolerable daily intake (PTDI) of heavy metals from the consumption of spices, we could not compare the EDI with standard PTDI.

**Table 2.**
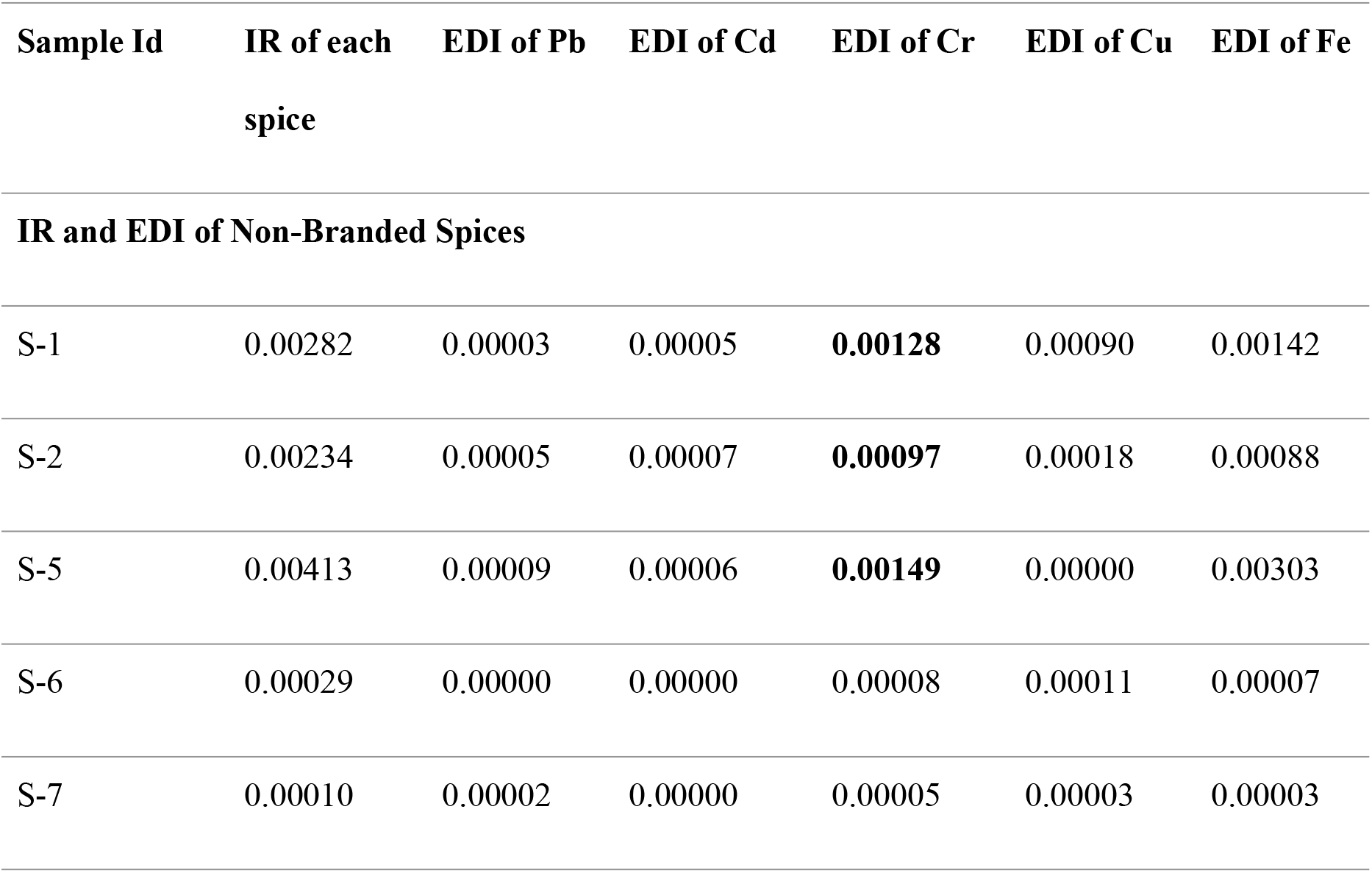
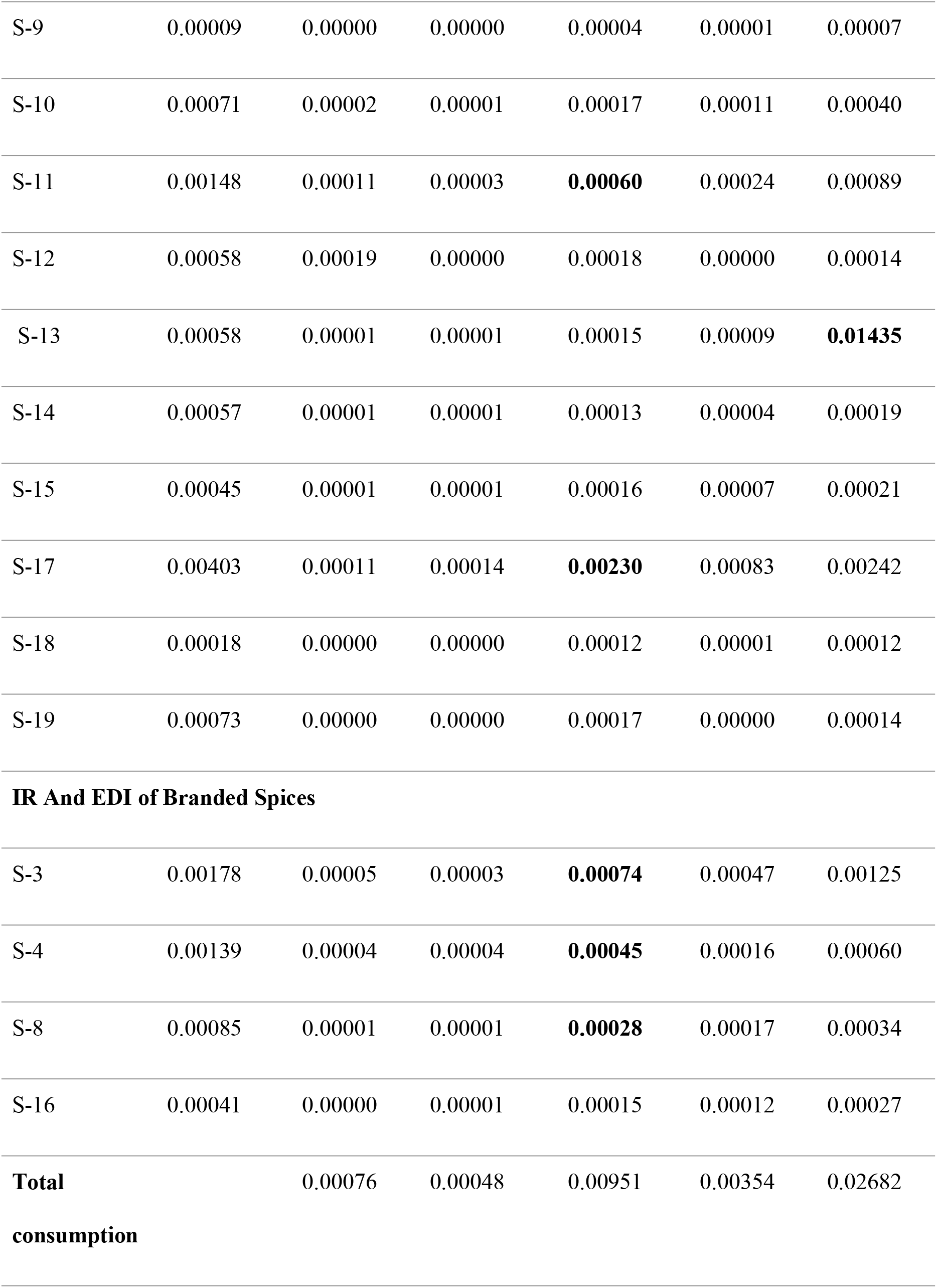
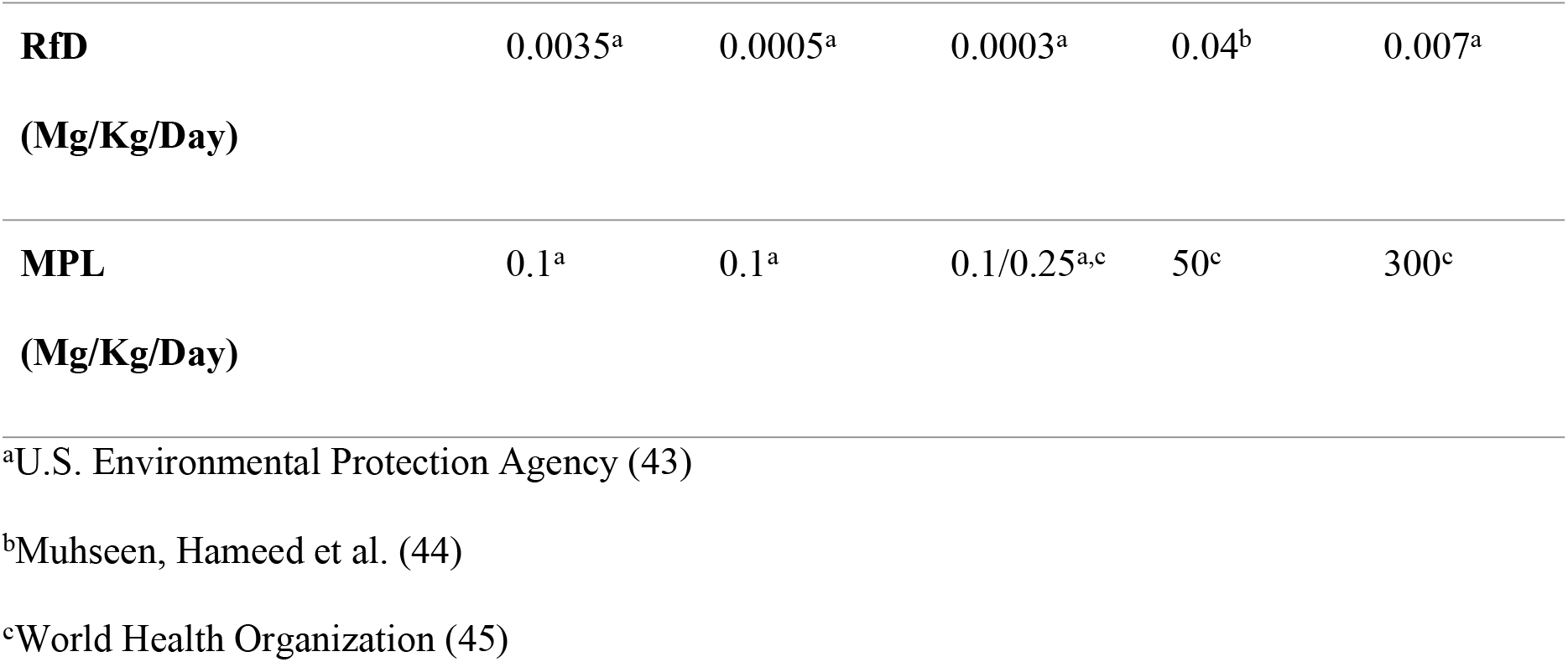
IR of spices (kg/day/person) and EDI of selected heavy metals through each sampled spice.

### Non-carcinogenic Risk for Selected Heavy Metals

To assess the risk of heavy metals in the Bangladeshi population, THQ and TTHQ were estimated and presented in Table 3. All THQ values except Cr for chili powder (NB), turmeric powder (NB), chili powder (B), turmeric powder (B), green chili, ginger, and coriander leaf were lower than 1, indicating low health risk. The mean THQ value for Pb was found to be lower than those from Nigeria (0.125) and Pakistan (0.281) (9, 22). The TTHQ values in the studied spices ranged from 0.135 to 8.328. The TTHQ values in some spices were >1, indicating potential health risk. Extremely high TTHQ values of Cr have been found in Green Chili, ginger, coriander leaf, and all forms of chili powder and turmeric powder. Our research had a higher TTHQ score than previous studies conducted in Poland, Pakistan, Egypt, and Nigeria (9, 22, 46, 47).

**Table 3:**
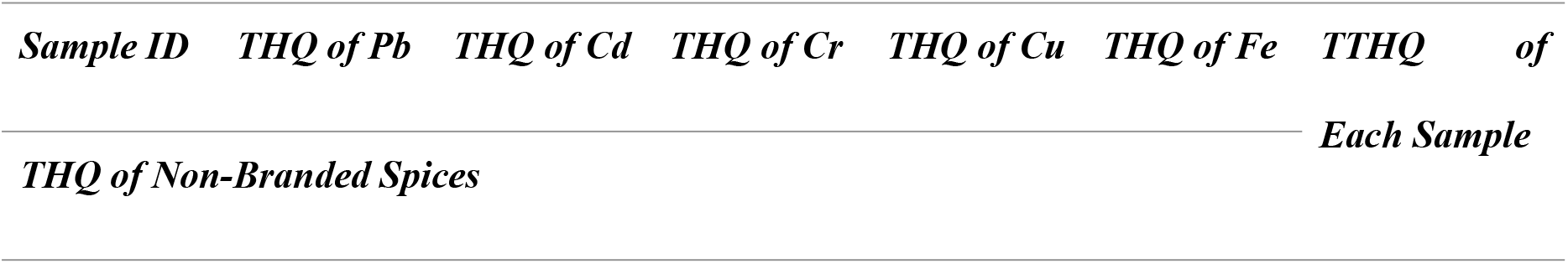
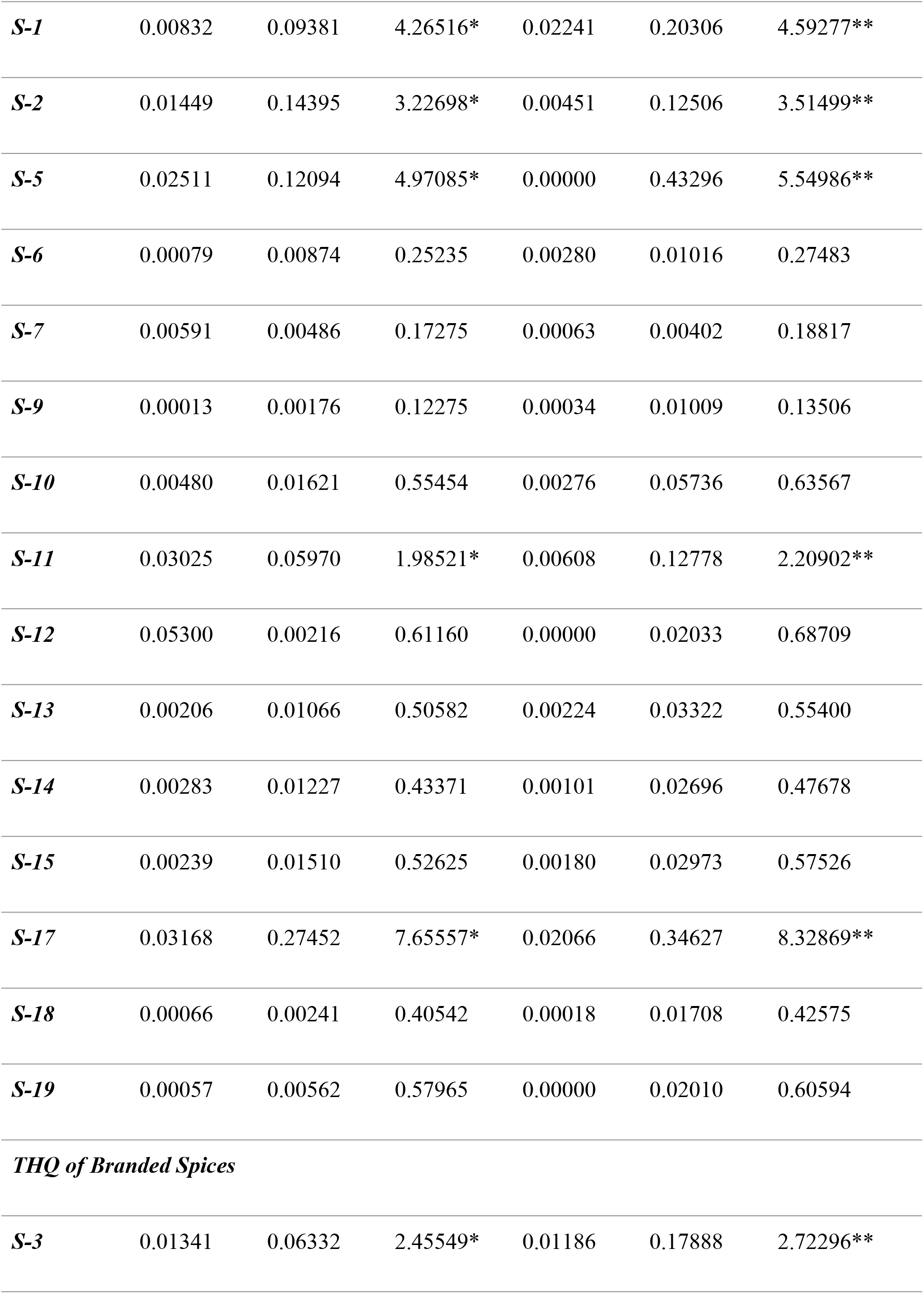
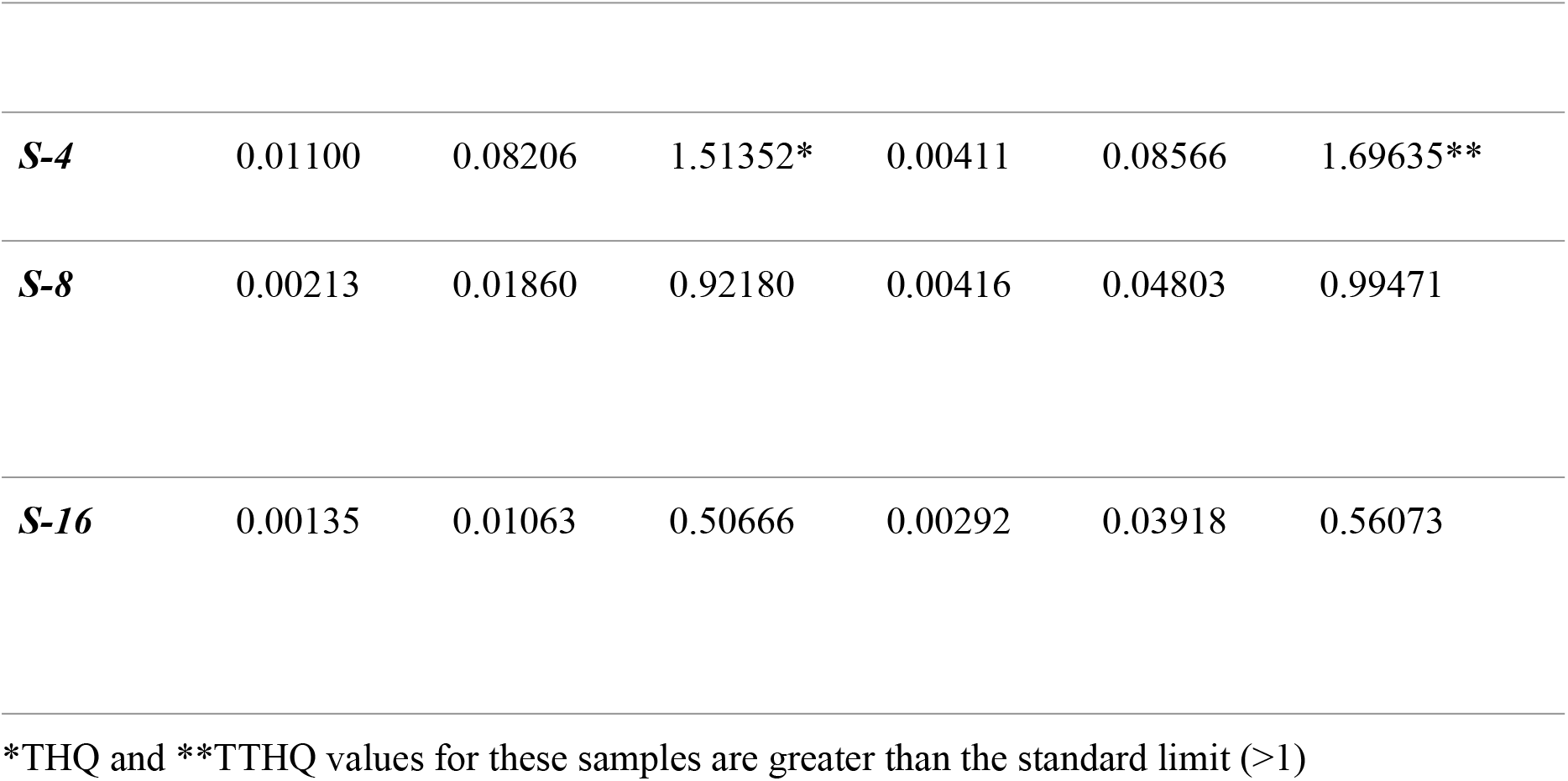
THQ and TTHQ of heavy metals through consumption of each sampled spice.

### Cancer Risk (CR) of Selected Heavy Metals

Figure 3 shows the total cancer risk of heavy metals in spices commonly found in local Bangladeshi markets. The CR of Cr and Cd for both branded and unbranded turmeric and chili powder, green chili, masala mix, ginger, and coriander leaf were higher than 1E-04, indicating that these spices were not safe for human consumption. Following the analysis, the non-branded chili and turmeric powder, green chili, and coriander leaf have the highest cancer risk of all samples in terms of chromium and cadmium.

**Figure 3.**
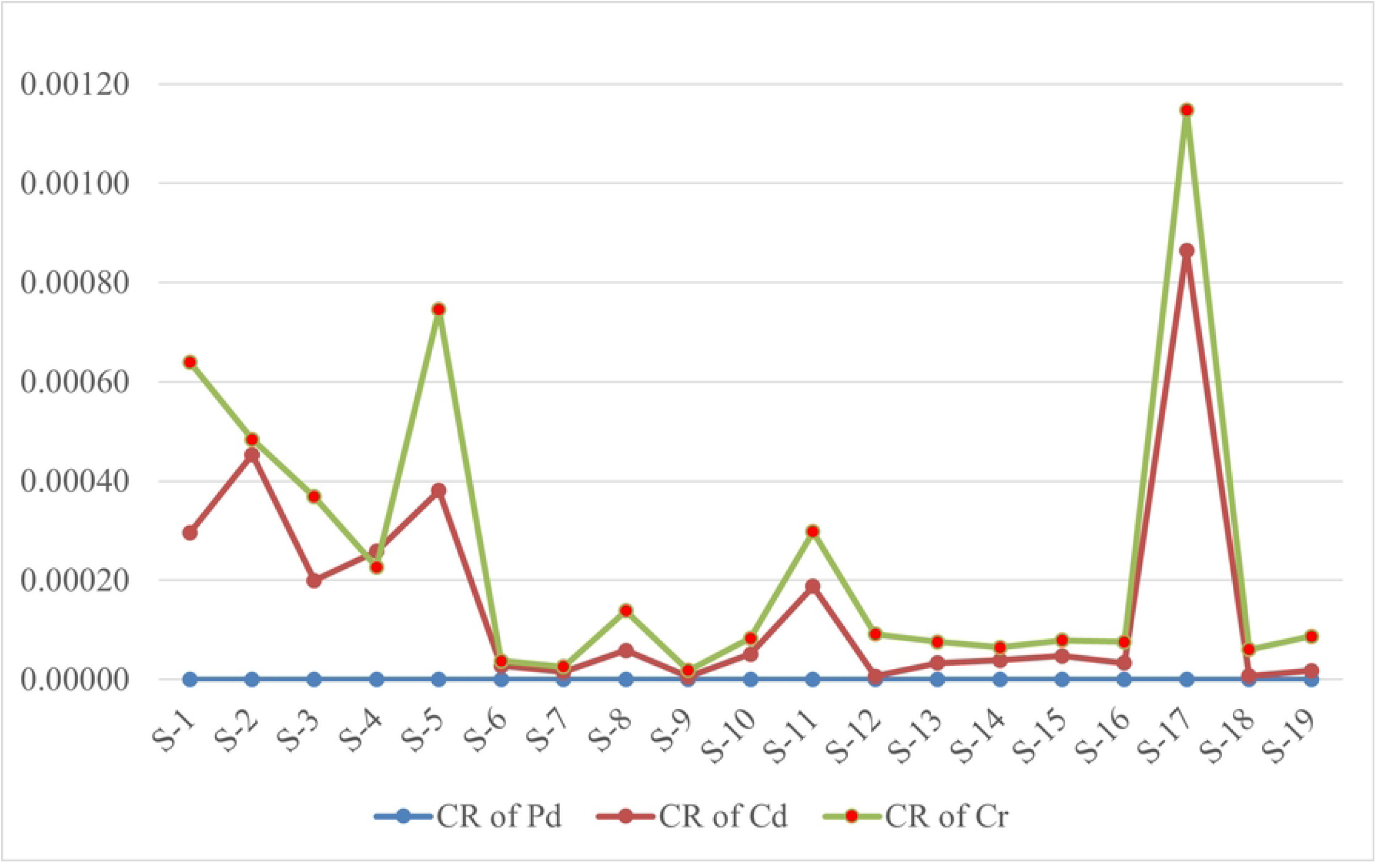
Cancer risk (CR) of heavy metals through consumption of each sampled spice

Our current study has certain limitations. We have analyzed a limited number of samples that were available at that specific point in time. Our study only presents the amount of specific heavy metal contents present in our sample but does provide information on the source of contamination. Furthermore, the ingestion rate of the households was computed based on each household’s daily or weekly spice usage.

## Conclusion

This research will assist regulatory organizations in increasing and improving heavy metal monitoring and evaluation procedures in spices, as well as ensure public health safety in relation to these food commodities. However, an extensive study is necessary to detect more elements in all consumed spices as well as the growing environment and supply chain, such as agricultural soil, water, packaging, etc., to find the source of contamination using a large sample size that represents the regional consumption patterns.

## Data Availability

All relevant data are within the manuscript and its Supporting Information files.

## Acknowledgement

We acknowledge University Grants Commission of Bangladesh for funding this research through UGC research grant 2020-2021.

## Notes

### Competing Interest Statement

The authors have declared no competing interest.

### Funding Statement

MRA received UGC research grant 2020-2021 (Grant no: Biological Science-2020/21-75) from University Grants Commission of Bangladesh (https://ugc.portal.gov.bd/). The funders had no role in study design, data collection and analysis, decision to publish, or preparation of the manuscript.

### Author Declarations

Ethical permission was obtained from the institutional ethics committee of the Noakhali Science and Technology University

